# Genetic and phenotype analyses of primary lateral sclerosis datasets from international cohorts

**DOI:** 10.1101/2023.07.19.23292817

**Authors:** Munishikha Kalia, Thomas P. Spargo, Ahmad Al Khleifat, Sarah Opie-Martin, Renata Kabiljo, Richard JB Dobson, Philip van Damme, Philippe Corcia, Philippe Couratier, Orla Hardiman, Russell McLaughlin, Marc Gotkine, Vivian Drory, Vincenzo Silani, Nicola Ticozzi, Jan H. Veldink, Leonard H. van den Berg, Mamede de Carvalho, Susana Pinto, Jesus S. Mora Pardina, Monica Povedano, Peter M. Andersen, Markus Weber, Nazli A. Başak, Christopher E Shaw, Pamela J. Shaw, Karen E. Morrison, John E. Landers, Jonathan D. Glass, Patrick Vourc’h, Project MinE ALS Sequencing Consortium, Ammar Al-Chalabi, Alfredo Iacoangeli

## Abstract

Primary lateral sclerosis (PLS) is the rarest form of motor neurone disease (MND). It is characterized by upper motor neuron degeneration, leading to progressive weakness, spasticity and functional disability. Although PLS does not typically shorten life substantially, it gradually impacts quality of life as the diseases progresses. There is no established genetic cause of PLS. One of the biggest challenges faced by people with PLS is delayed diagnosis and misdiagnosis, since the initial symptoms can be similar to amyotrophic lateral sclerosis (ALS), the most common form of MND. In the absence of a concrete genetic test that differentiates PLS from other MNDs, this delay in diagnosis is inevitable. Understanding the genetic basis of PLS might help in reducing the time from the onset of symptoms to diagnosis, and it will improve our understanding of the disease biology favouring the development of a treatment.

The aim of our study is to collect a large international PLS genetic and clinical dataset to investigate its genetic and phenotypic landscapes as well as to evaluate whether genetic testing should be advised in PLS. Through Project MinE and AnswerALS, we accessed whole-genome sequencing data of 120 PLS, 7405 ALS and 2444 controls. We identified variants in several MND genes such as *FIG4, FUS, SPG7, SPG11* and *SQSTM1* genes among others and repeat expansions in the *ATXN1* (12.2%) and *NIPA1* (7.3%) genes, but none in the *C9orf72* and *ATXN2* genes. Overall PLS patients harboured fewer clinically actionable MND-associated variants than ALS patients (p = 0.0001), however, depending on the panel, up to 11% of people with PLS might benefit from genetic testing. By looking at the clinical characteristics of these cohorts, the age of symptom onset was not younger for people with PLS than for those with ALS in both Project MinE and AnswerALS. On such bases, we advise that the current diagnostic criteria that discourage the use of genetic testing and rely on age of onset should be reconsidered.

## INTRODUCTION

Primary lateral sclerosis (PLS) is a slowly progressive neurodegenerative disease characterized by pure upper motor neuron (UMN) dysfunction for at least the first four years from onset [1]. The lack of lower motor neuron (LMN) involvement distinguishes PLS from amyotrophic lateral sclerosis (ALS) [2]. PLS is considered to be the rarest form of motor neurone disease (MND), representing approximately 1-4 % of all MNDs [2-7]. For the majority of people with PLS, the symptoms originate in the lower extremities, reporting progressive weakness and stiffness of the legs, leading to an insidious loss of mobility [6]. The symptoms less commonly emerge in the corticobulbar pathways, usually manifesting as dysarthria, followed by dysphagia [6]. Akin to ALS, PLS also seems to have a male predominance [6]. Although the motor function is immensely impaired in PLS, it is not fatal like ALS. PLS mostly develops in midlife, and the mean age of onset of PLS reported in literature is around 50 years, a decade earlier than ALS [1, 6]. However, a juvenile form exists, which occurs in childhood and young adults and is caused by mutations in the *ALS2* gene [8].

Due to the rarity of PLS, there are no large-scale genetic studies to identify genes associated with PLS, therefore, the genetic architecture of the adult form of PLS is not well known but is thought to be a heterogenous disease similar to ALS [2, 9, 10]. Few studies have been performed before and have implicated several MND genes with PLS [10-18]. In addition, *C9orf72* repeat expansions, the most common genetic causes of ALS, have only been reported in three people with PLS [10, 13, 18]. Whilst familial transmission has been observed in juvenile PLS, adult PLS is mostly sporadic, however, some recent studies have reported familial cases of PLS [11, 12, 16, 17].

Currently, the cause and pathogenic mechanism of PLS are mostly unknown and no treatments are available. Moreover, one of the biggest challenges faced by people with PLS is misdiagnosis and delayed diagnosis due to the overlap of symptoms with UMN dominant ALS. Understanding the genetic basis of PLS is crucial as it might help to reduce the diagnosis time from the onset of symptoms, and most importantly, by improving our understanding of its biological causes, it might lead to the development of treatments.

In order to be able to investigate the genetics of PLS we have collected whole-genome sequencing (WGS) data of 120 PLS, 7405 ALS and 2444 control samples from the Project Mine [19] and AnswerALS [20] datasets. Here we report the findings from our screening of MND-linked genes in these samples and the results of our phenotype characterisation of the PLS cohorts with respect the ALS cohorts from the same datasets.

## RESULTS

### Demographic and clinical characterisation of the cohorts

We acquired access to whole-genome sequencing (WGS) and clinical data of the PLS and ALS patients, and 2444 controls through the Project Mine [19] and AnswerALS [20, 21] international initiatives. The demographic and clinical details of the cohorts are presented in Table 1. All the people with PLS in this study did not have a family history of PLS.

**Table 1.** Cohort demographics and phenotypes. *Frontotemporal dementia.

|  |  | Project MinE | AnswerALS |
| --- | --- | --- | --- |
| Dataset | PLS | 81 | 38 |
|  | PLS-FTD* | 1 | - |
|  | ALS | 6550 | 855 |
|  | Controls | 2444 | - |
| PLS diagnosis | Definite | 82 | - |
|  | Suspected | - | 19 |
|  | Possible | - | 11 |
|  | Probable | - | 1 |
| Sex | Male | 41 | 22 |
|  | Female | 41 | 16 |
| Site of onset | Bulbar | 62 | 9 |
|  | Spinal | 10 | 28 |
|  | Generalized | 8 | 1 |
| Mean age of onset (years) | PLS | 59.3 | 56.4 |
|  | ALS | 60.1 | 56.5 |
| Mean diagnostic delay (years) | PLS | 3.5 | - |
|  | ALS | 1.26 | - |

### Genetic screening of MND genes

In Table 2 and 3 we report all rare variants with high and moderate functional effect found in the PLS patients in 164 MND-linked genes. We identified known pathogenic variants (ClinVar) in PLS patients: *FUS, FIG4, SPG7, SPG11* and *SQSTM1* genes (Table 2), none of these variants were identified in our control population. The person with the *SPG11* (p.Arg651X) variant had PLS-FTD diagnosis. In addition to the pathogenic variants, a novel variant was identified in the *TBK1* (p.T682Hfs*2) gene in a PLS patient. A novel missense variant was identified in *ATXN2*, which was predicted deleterious by SIFT and a novel frameshift variant was identified in *TBK1*. We identified two previously characterized pathogenic variants in the *PNPLA6* and *LIPC* genes, which were present both in our PLS cohort and the control population (Table 3).

**Table 2.** List of high and moderate impact variants identified in the PLS cohort but not in controls. All the variants listed below were present in the heterozygous form in the PLS patients. SETX^**@**^: same individual, VUS* variant of uncertain significance, M^$^: Patient with PLS-FTD diagnosis, SPG7^&^: Patient misdiagnosed with PLS (see discussion).

| Gene name | Sex | Censored | Disease duration (years) | Site of onset | Cohort | Variant | Protein change | Consequence | CADD-PHRED | SIFT | ClinVar | Frequency gnomAD | Frequency in PLS cohort (%) |
| --- | --- | --- | --- | --- | --- | --- | --- | --- | --- | --- | --- | --- | --- |
| *ALS2* | M | Y | 5.9 | Spinal | SP | c.1280C>A | p.Thr427Lys | Missense | 19.1 | Tolerated | - | 8.84e-6 | 0.83 |
| *ATXN2* | M | N | 12.3 | Bulbar | UK | c.1322C>T | p.Tyr441Met | Missense | 26.3 | Deleterious | - | - | 0.83 |
| *C9orf72* | M | Y | 6.4 | Spinal | US | c.971A>G | p.His324Arg | Missense | 25.9 | Deleterious | - | 8.82e-6 | 0.83 |
| *ERBB4* | F | Y | 10.0 | Spinal | UK | c.1829C>G | p.Pro610Arg | Missense | 16.8 | Tolerated | - | 3.52e-5 | 0.83 |
| *FIG4* | F | N | 3.5 | Generalized | UK | c.1373dup | p.Leu458Ffs*5 | Frameshift | - | - | - | 4.40e-5 | 0.83 |
| *FIG4* | M | Y | 6.6 | Spinal | IT | c.68G>A | p.Arg23Lys | Missense | 24.8 | Tolerated | Pathogenic | - | 0.83 |
| *FUS* | M | N | 5.5 | Spinal | NL | c.1561C>T | p.Arg521Cys | Missense | 24.1 | Deleterious | Pathogenic | 2.64e-5 | 0.83 |
| *MATR3* | M | - | - | Spinal | FR | c.1640A>G | p.Asn547Ser | Missense | 20.9 | Tolerated | VUS* | 2.74e-4 | 0.83 |
| *SETX*<sup>®</sup> | F | N | 5.3 | Spinal | UK | c.5084A>C | p.Gln1695Pro | Missense | 25.0 | Deleterious | - | - | 0.83 |
| *SETX*<sup>®</sup> | F | N | 5.3 | Spinal | UK | c.3311A>C | p.Gln1104Pro | Missense | 16.1 | Deleterious | - | - | 0.83 |
| *SPG7*<sup>&</sup> | M | Y | 19.8 | Spinal | US | c.233T>A | p.Leu78X | Stop gain | 33.0 | - | Pathogenic | 12.3e-6 | 0.83 |
| *SPG7* | M | - | - | Bulbar | US | c.1529C>T | p.Ala510Val | Missense |  | Deleterious | Likely Pathogenic | 35.9e-5 | 0.83 |
| *SPG11* | M<sup>§</sup> | Y | 8.5 | Spinal | BE | c.1951C>T | p.Arg651X | Stop gain | 36.0 | - | Pathogenic | 4.41e-5 | 0.83 |
| *SQSTM1* | F | N | 6.0 | Spinal | US | c.C1175C>T | p.Pro392Leu | Missense | 25.2 | Deleterious | Pathogenic | 13.3e-5 | 0.83 |
| *TARDBP* | M | Y | 4.9 | Spinal | UK | c.71G>A | p.Gly24Asp | Missense | 25.6 | Deleterious | - | - | 0.83 |
| *TBK1* | F | Y | 2.1 | Spinal | PT | c.2044delA | p.T682Hfs*2 | Frameshift | - | - | - | - | 0.83 |

**Table 3.**
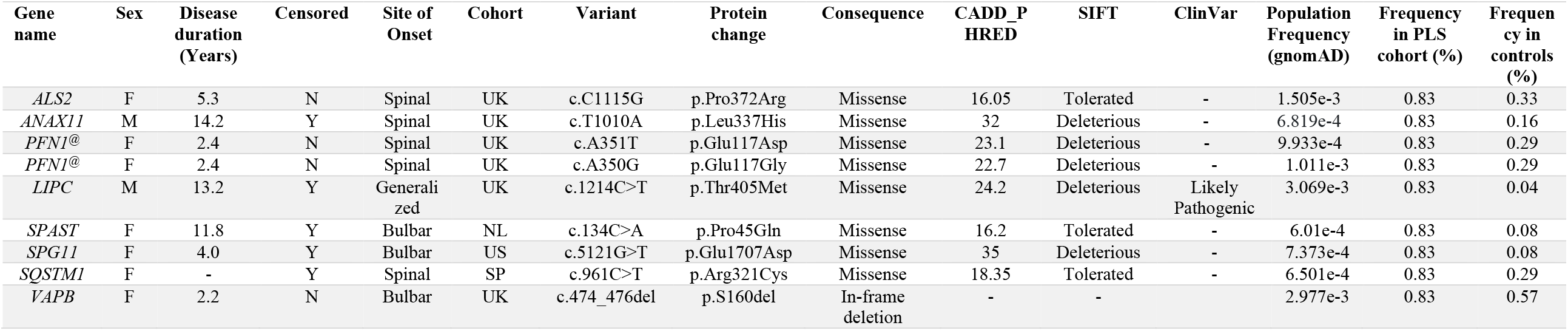
High and moderate impact variants identified both in PLS patients and control population. All the variants listed below were present in a heterozygous form in the PLS patients and controls. PNF1^**@**^: same individual

### Pathogenic repeat expansions in PLS

Pathogenic repeat expansions in *ATXN1, ATXN2, C9orf72 & NIPA1* genes are associated with ALS [22-27]. Two previous studies report presence of the *C9orf72* repeat expansion in a PLS patient [10, 13]. However, the repeat expansions in *ATXN1, ATXN2 & NIPA1* have not been found in people with PLS before. Here, we studied the presence of repeat expansions in the PLS dataset. A total of 120 patients were sampled for the *ATXN2* and *C9orf72* repeat expansions and 82 patients for the *ATXN1* and *NIPA1* repeat expansions (Table 4). No pathogenic *ATXN2* and *C9orf72* repeat expansions were observed in our dataset. We found ALS-associated repeat expansions in *ATXN1* (12.2 %) and *NIPA1* (7.3%) in the PLS patients.

**Table 4.** Pathogenic repeat expansions in PLS.

| Gene name | Repeat expansions | No. of samples | Repeat length in normal population | Pathogenic expansion in ALS | Expected frequency ALS (%) | Frequency observed PLS (%) |
| --- | --- | --- | --- | --- | --- | --- |
| *ATXN1* | CAG/CAT | 82 | <29 | ≥39 | 12.0 | 12.2 |
| *ATXN2* | CAG | 120 | 22-23 | 29-33 | 1.3 | 0 |
| *C9orf72* | G4C2 | 120 | 2 | >30 | 8.0 | 0 |
| *NIPA1* | GCG | 82 | <7 | >8 | 5.54 | 7.3 |

### Comparison of PLS and ALS genetic screening

In order to assess whether people with PLS would benefit from genetic screening using MND gene panels, we compared the proportion of patients that would present potential clinically actionable variants between the PLS and ALS cohorts [28]. For this analysis we drew upon the recently published ALS gene panels and definition of “potentially clinical actionable variants” (see Methods) [28]. The 3 panels were: i) a four-gene panel with the most common MND genes that included pathogenic variants in *FUS, TARDBP, SOD1* and the *C9orf72* repeat expansion, ii) a “Large” panel, consisting of 24 genes selected for harbouring likely large-effect, rare Mendelian ALS gene variants, and iii) a panel consisting of 26 genes selected for harbouring likely large-effect, rare Mendelian ALS gene variants from Genomics England [29]. Only 2 PLS patients were positive when tested with the 4-gene panel (the *FUS* and *TARDBP* carriers), 6% with the Large panel (7 out of 119), and approximately 11% (13 out of 119) were positive with the Genomics England panel (Figure 1). Across all gene panels, variants in screened genes were less frequent among people with PLS than those with ALS; respective to each panel, variants occurred in approximately 8%, 20% and 21% of the ALS cohort (p_4-genes_ = 0.0057, p_24-genes_ = 0.0001 and p_26-genes_ = 0.0064, Figure 1). Clinical interpretation of the variants found in the PLS patients are available in Supplementary Table 1 and 2.

**Figure 1.**
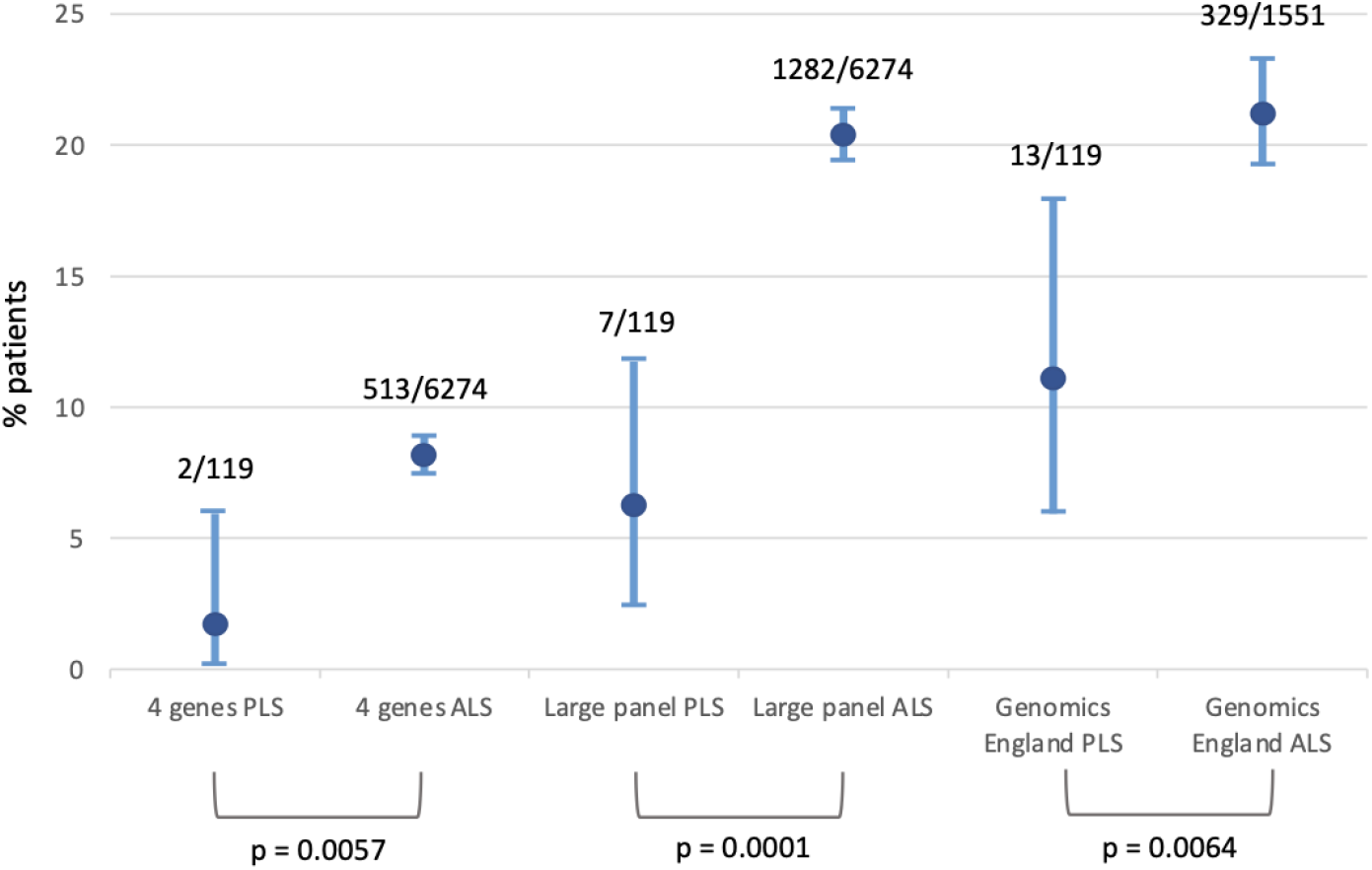
Percentage of people with PLS and ALS with a clinically actionable variant using the 4-genes, Large and Genomics England panels. 6274 ALS patients were tested for the 4-gene and 24-gene Large gene panel, 1551 British ALS patients were tested for the Genomics England panel. 119 PLS patients were tested for all panels. Error bars denote 95% CI.

### Phenotype analysis

Phenotype analysis was performed on the PLS and ALS samples with relevant clinical data available from the AnswerALS and Project Mine initiatives. Kaplan–Meier analysis was performed to compare the age of symptom onset (Figure 2) and survival from symptom onset until death or censoring (Figure 3) between PLS and ALS patients in the Project MinE and AnswerALS datasets; differences were tested using pairwise log-rank tests. For age of onset analysis, no significant differences were observed between the PLS and ALS cohorts from either dataset (log-rank test p>0.05). The mean age of onset in the Project MinE PLS and ALS patients was 59.3 and 60.1 years respectively. For AnswerALS, the mean age of onset was 56.4 and 56.5 years in PLS and ALS respectively. Next, we compared survival from symptom onset for ALS and PLS patients. As expected, the survival of ALS and PLS (Project MinE) patients was highly different (p<0.0001) with the estimated medians equal to 3.02 and 16.82 years in ALS and PLS patients respectively. There was no survival data available for the PLS patients from AnswerALS, therefore, only the Project MinE dataset was utilized for this comparison (Table 5).

**Table 5.** Summary table of the Kaplan–Meier survival analysis.

|  | Dataset | Phenotype | N | Events | Rmean<br>(years) | StdErr<br>(years) | Median<br>(years) | 0.95LCL<br>(years) | 0.95UCL<br>(years) |
| --- | --- | --- | --- | --- | --- | --- | --- | --- | --- |
| Age of Onset | Project MinE | ALS | 6316 | 6316 | 60.1 | 0.16 | 61.5 | 61.2 | 61.9 |
|  |  | PLS | 79 | 79 | 59.3 | 1.24 | 59.0 | 56.0 | 63.3 |
|  | AnswerALS | ALS | 832 | 832 | 56.5 | 0.398 | 57 | 57 | 58 |
|  |  | PLS | 38 | 38 | 56.4 | 1.480 | 56 | 54.0 | 61.0 |
| Survival | Project MinE | ALS | 5285 | 4330 | 1.79 | 0.0856 | 0.991 | 0.961 | 0.999 |
|  |  | PLS | 74 | 25 | 14.39 | 3.3682 | 8.000 | 5.002 | NA |

**Figure 2.**
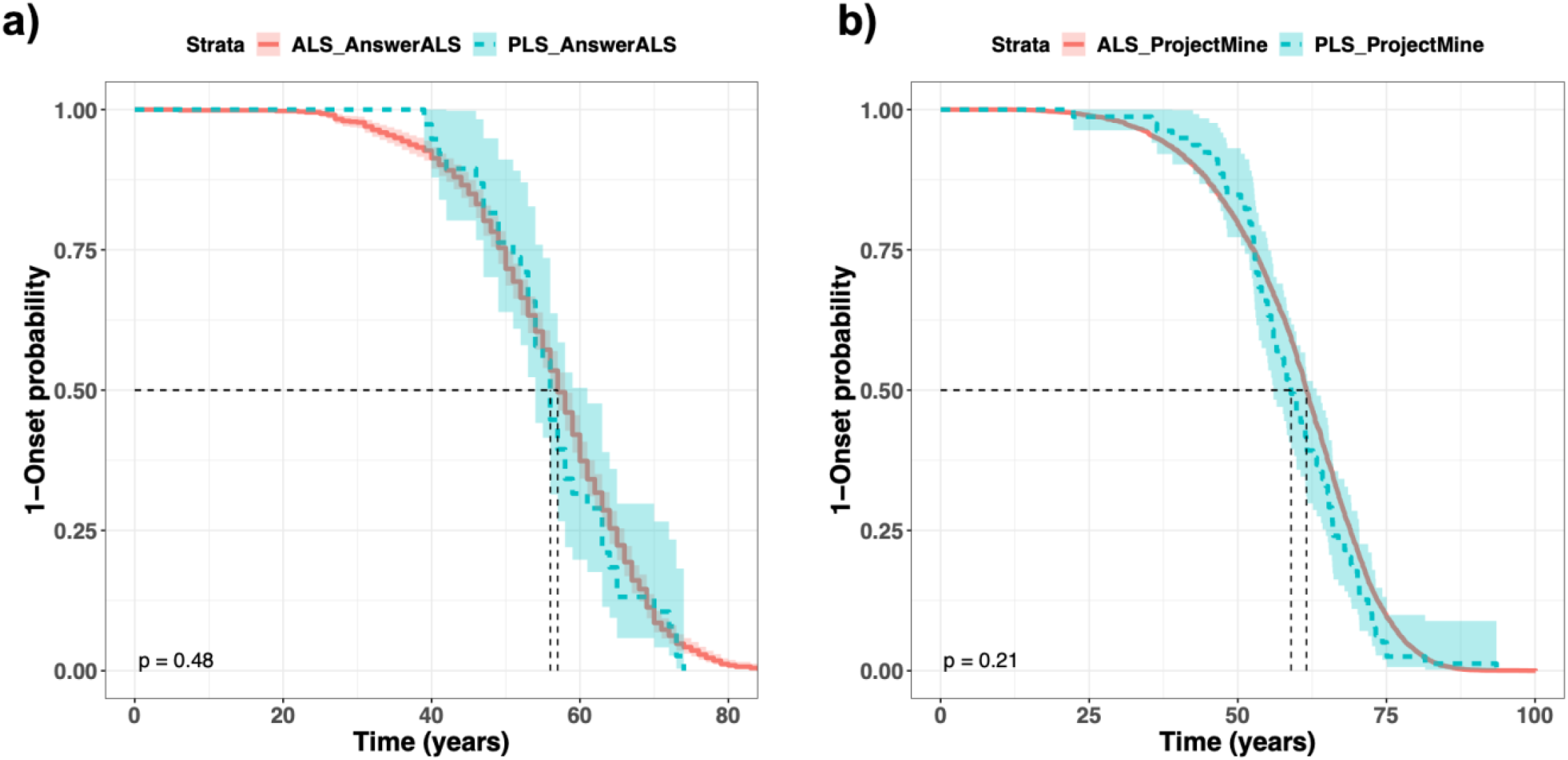
Kaplan–Meier curves for the age of onset comparison in ALS & PLS patients from the a) AnswerALS and the b) Project MinE datasets.

**Figure 3.**
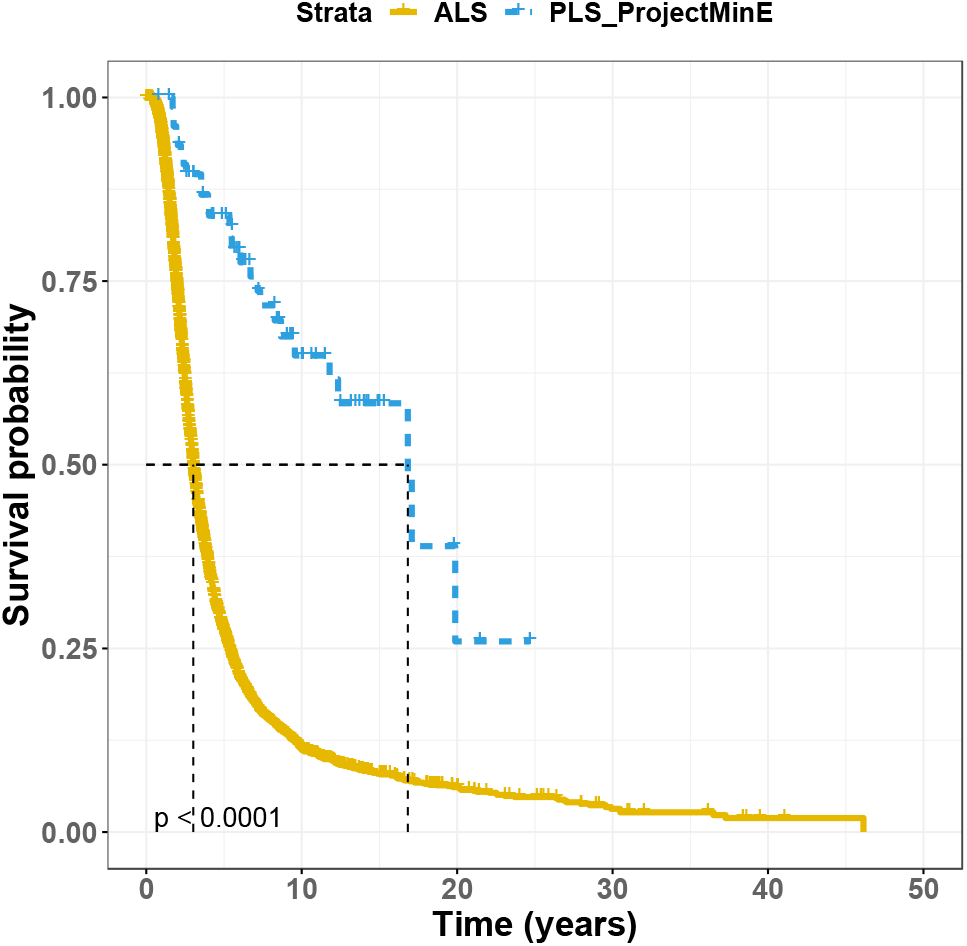
Survival probability comparison between the ALS and PLS patients in the Project Mine dataset.

## DISCUSSION

In this study we report the results of the genetic screening and phenotype analysis from large cohorts of PLS patients obtained from the Project MinE and AnswerALS international consortia. We identified people with PLS harbouring known pathogenic variants in the *FUS, FIG4, SQSTM1, SPG7* and *SPG11* genes. The *SPG7* mutation (p.Leu78Ter) has been previously associated with an early onset of hereditary spastic paraplegia (HSP) [30, 31]. The *SPG11* mutation (p.Arg651Ter) has also been previously associated with HSP [32, 33]. Since these variants are involved in the HSP and misdiagnosis is a common issue in PLS, we contacted the clinicians involved in the diagnosis of these two patients. The clinicians reported that the patient with *SPG7* variant had been misdiagnosed with PLS, and they most likely suffered with HSP. Whilst the patient with the *SPG11* was a definite PLS patient with frontotemporal dementia. In a different PLS patient, we found another pathogenic *SPG7* (p.Arg510Val) gene variant in heterozygous form. This variant has been reported previously in one PLS patient [10] and, in homozygous form, is also the most commonly reported variant responsible for the adult onset HSP in people of British ancestry [34]. The variant identified in the *FUS* gene (p.Arg521Cys) has been previously associated with ALS and characterized by rapid progression and high penetrance [35-37]. The pathogenic variant in *FIG4* (p.Leu458Ffs*5) has been associated with Charcot-Marie-Tooth disease [38]. The variant in *SQSTM1* has been previously associated with FTD-ALS in the French population [38].

We identified pathogenic repeat expansions in *ATXN1* and *NIPA1*genes in our PLS cohort. The frequency of *ATXN1* repeat expansions in our cohort was similar to that observed previously in ALS [22]. However, the frequency of *NIPA1* in our PLS cohort was slightly higher than that observed in ALS studies [27]. These expansions have not been reported before in PLS patients. Our investigation of pathogenic repeat expansions in *ATXN2* and *C9orf72* genes showed no expansions in the PLS cohort. It is important to note here that pathogenic repeat expansions in *C9orf72* gene have only been reported in three PLS patients in previous studies [10, 13, 18] and no other repeat expansions have ever been reported. Our observations suggest that these expansions do not play a vital role in PLS, in contrast to ALS.

We investigated the use of genetic screening in PLS using MND gene panels. Our results showed that across the three panels tested, PLS patients presented a significantly lower number of clinically actionable variants with respect to ALS patients. However, between 2% to 11% of people at risk of developing PLS, depending on the panel, might benefit from genetic testing. These results are consistent with a recent large-scale Dutch study that showed that approximately 7% of the 139 people with PLS they screened harboured a pathogenic or likely pathogenic variant. Together, these results strongly support the use of genetic testing for the diagnosis of PLS.

Finally, the phenotype analysis showed the age of onset in the PLS patients from the Project MinE and AnswerALS cohorts was 59.3 and 56.3 years respectively and negligible differences were observed with respect to the corresponding ALS cohorts. This is one key findings of our study, as currently the age of onset in PLS is thought to be approximately 50 years, 10 years younger than in ALS, and it is a contributing factor to the diagnosis [1, 6]. Our results suggest this might need to be reconsidered.

Overall, using a large dataset of PLS we were able to assess the genetic and phenotype landscape of PLS, and to investigate the utility of genetic testing in PLS. Our results set the basis for a large-scale approach to the study of PLS which, although highly challenging given the rarity of the disease, is essential to elucidate its genetic causes.

## METHODS

### Whole genome sequencing

The detail methods employed in the whole genome sequencing for the Project Mine dataset are described here [19]. Briefly, DNA was extracted from the venous blood drawn from patients and controls. Illumina’s FastTrack services (San Diego, CA, USA) on the Illumina HiSeq 2000 platform were used for DNA sequencing. PCR free library preparation was used for sequencing, which was 100 bp paired end and yielded ∼40X coverage across each sample. The sequences were aligned to hg19 reference genome using Isaac pipeline and to call single nucleotide variants [39]. Variants which did not pass the Isaac filtering criteria were set to missing, non-autosomal chromosome and multi-allelic variants were excluded. PLINK [40, 41] and VCFtools [42] software were used to perform sample and SNP quality control.

For the AnswerALS [21] whole-genome sequencing DNA was extracted from the whole blood or the flash frozen post-mortem tissue. The libraries were prepared using the Illumina TruSeq DNA PCR-free Library Preparation Kit. The sequencing data was processed on an NYGC automated pipeline. Paired end reads were aligned to the GRCh38 human reference genome by means of Burrows-Wheeler Aligner [43] and the processing was done by following the GATK best practice workflow [44]. Picard tools (http://broadinstitute.github.io/picard/) was used to mark the duplicate reads, local realignment around indels and base quality score recalibration was done via Genome Analysis Toolkit [45].

### Variant annotation and prioritization

Variant annotation was performed with ANNOVAR [46] and the Ensemble-VEP [47] software. We tested for variants in 154 candidate genes obtained from ALSoD database (list available at https://alsod.ac.uk) [48] and the additional genes *ERLIN2, MMACHC, SYNE2, TTBK2, D4S2963, PARK2, CLN6, BTD, LRKK2, KIF1A* identified from literature review of the previous PLS studies [10]. The variants were filtered on the basis of allele frequency information from the gnomAD database and functional effect. Variants with minor allele frequency (MAF) < 0.005 and with a predicted moderate and high functional impact on gene function as defined by VEP, were selected. In brief, using consequence terms from the sequence ontology, moderate impact variants included missense, in-frame insertions and deletions and protein altering variants. High impact variants included stop lost and gained, start lost, transcript amplification, frameshift, transcript ablation and splice acceptor and donor variants. The genome coordinates of the variants aligned to the human reference genome hg19 were converted to the human reference genome hg38 using LiftOver [49].

### Panel screening

Genetic screening of PLS and ALS cohorts in this study followed our recently published protocol [28]. Briefly, three gene panels were used. The first included the *C9orf72* repeat expansion, all missense rare variants in *SOD1*, and *FUS* and *TARDBP* reported to be pathogenic or likely pathogenic in the ClinVar database or present in AlSoD with a linked publication and found in 2 or more patients. The second Large panel, consisted of 24 genes selected for harbouring likely large-effect, rare Mendelian ALS gene variants, and a third panel included clinically actionable variants in 26 genes from the Genomics England ‘Amyotrophic lateral sclerosis/motor neurone disease v1.48’ panel. The genes included in the panels have been previously described [28]. Clinical actionable variants were defined as those classified pathogenic, likely pathogenic or VUS with high probability of pathogenicity by our automated ACMG pipeline [28]. All PLS cases, 119 after removing the *SPG7* patient who most likely had HSP, were included in this analysis. The results of the screening in ALS patients from Project MinE from our previous work [28] were used. These included 6274 ALS patients tested for the 24-gene Large gene panel and 1551 British ALS patients tested for the Genomics England panel. Fisher’s exact test was used to test the differences between ALS and PLS results.

### *ATXN1* expansion testing

CAG/CAT repeats were tested in *ATXN1* gene. Expansions <29 are considered normal, whilst expansions >33 are considered pathogenic in ALS and expansions ≥39 are considered pathogenic in Spinocerebellar Ataxia type 1.

### *ATXN2* expansion testing

Polyglutamine expansions GCG repeats were tested in the PLS cohort. Expansions <29 are considered normal, whilst expansions 29-33 are considered pathogenic in ALS and expansions >34 are considered pathogenic in Spinocerebellar Ataxia type 2.

### *C9orf72* expansion testing

The *C9orf72* hexanucleotide repeat expansions were tested. Expansions <30 are considered normal and expansions ≥30 are considered pathogenic in ALS [50]. We used Expansion Hunter and data obtained from real time PCR to confirm *C9orf72* expansion status. Following Expansion Hunter tool instructions, the genome coordinates that were used to confirm *C9orf72* expansion status were chr9:27573527-27573544 and the motif GGCCCC [50]. Additionally, 29 off-target regions were also included to determine the *C9orf72* repeat size. Please refer to the Expansion Hunter Github page (https://github.com/Illumina/ExpansionHunter) for the exact coordinates of the 29 off-target regions).

### *NIPA* expansion testing

Polyalanine expansions CAG repeats were tested in the PLS cohort. Expansions <7 are considered normal and >8 are considered pathogenic in ALS [51].

### Survival Analysis

Kaplan–Meier method was used to estimate the survival probability of ALS and PLS patients from the survival times [51]. Differences between survival curves were tested using the log-rank test. ‘survival’ [52] and ‘survminer’ [53] packages were utilized from the statistical language R (https://cran.r-project.org) [54].

## Data Availability

All data produced in the present work are contained in the manuscript

## ACKNOWLEDGEMENTS

The authors thank all the study participants, their families, and carers. “Data used in the preparation of this article were obtained from the ANSWER ALS Data Portal (AALS-01184). For up-to-date information on the study, visit https://dataportal.answerals.org.”

We also acknowledge use of the research computing facility at King’s College London, Rosalind (https://rosalind.kcl.ac.uk), which is delivered in partnership with the National Institute for Health and Care Research (NIHR) Biomedical Research Centres at South London & Maudsley and Guy’s & St. Thomas’ NHS Foundation Trusts and part-funded by capital equipment grants from the Maudsley Charity (award 980) and Guy’s and St Thomas’ Charity (TR130505). The authors also acknowledge the use of the King’s Computational Research, Engineering and Technology Environment (CREATE) 2022. Available from: https://doi.org/10.18742/rnvf-m076.

The views expressed are those of the author(s) and not necessarily those of the NHS, the NIHR, King’s College London, or the Department of Health and Social Care.

## REFERENCES

1. Turner, M.R., et al., Primary lateral sclerosis: consensus diagnostic criteria. Journal of Neurology, Neurosurgery & Psychiatry, 2020. 91(4): p. 373–377.

2. Le Forestier, N., et al., Primary lateral sclerosis: further clarification. Journal of the neurological sciences, 2001. 185(2): p. 95–100.

3. Singer, M.A., et al., Primary lateral sclerosis: clinical and laboratory features in 25 patients. Journal of clinical neuromuscular disease, 2005. 7(1): p. 1–9.

4. Pringle, C., et al., Primary lateral sclerosis: clinical features, neuropathology and diagnostic criteria. Brain, 1992. 115(2): p. 495–520.

5. Barceló Rado, M.A., et al., Estimation of the prevalence and incidence of motor neuron diseases in two Spanish regions: Catalonia and Valencia. Scientific Reports, 2021, vol 11, art. núm. 6207, 2021.

6. Singer, M.A., et al., Primary lateral sclerosis. Muscle & Nerve: Official Journal of the American Association of Electrodiagnostic Medicine, 2007. 35(3): p. 291–302.

7. Statland, J.M., et al., Primary lateral sclerosis. Neurologic clinics, 2015. 33(4): p. 749–760.

8. Yang, Y., et al., The gene encoding alsin, a protein with three guanine-nucleotide exchange factor domains, is mutated in a form of recessive amyotrophic lateral sclerosis. Nature genetics, 2001. 29(2): p. 160–165.

9. Zhai, P., et al., Primary lateral sclerosis: A heterogeneous disorder composed of different subtypes? Neurology, 2003. 60(8): p. 1258–1265.

10. Mitsumoto, H., et al., Phenotypic and molecular analyses of primary lateral sclerosis. Neurology Genetics, 2015. 1(1).

11. Gómez-Tortosa, E., et al., Familial primary lateral sclerosis or dementia associated with Arg573Gly TBK1 mutation. Journal of Neurology, Neurosurgery & Psychiatry, 2017. 88(11): p. 996–997.

12. Hirsch-Reinshagen, V., et al., Clinicopathologic correlations in a family with a TBK1 mutation presenting as primary progressive aphasia and primary lateral sclerosis. Amyotrophic Lateral Sclerosis and Frontotemporal Degeneration, 2019. 20(7-8): p. 568–575.

13. Van Rheenen, W., et al., Hexanucleotide repeat expansions in C9ORF72 in the spectrum of motor neuron diseases. Neurology, 2012. 79(9): p. 878–882.

14. Chow, C.Y., et al., Deleterious variants of FIG4, a phosphoinositide phosphatase, in patients with ALS. The American Journal of Human Genetics, 2009. 84(1): p. 85–88.

15. Bergner, C.G., et al., Case Report: Association of a Variant of Unknown Significance in the FIG4 Gene With Frontotemporal Dementia and Slowly Progressing Motoneuron Disease: A Case Report Depicting Common Challenges in Clinical and Genetic Diagnostics of Rare Neuropsychiatric and Neurologic Disorders. Frontiers in Neuroscience, 2020. 14: p. 559670.

16. Liu, Y., et al., Exome sequencing identifies a mutation (Y740C) in spastic paraplegia 7 gene associated with adult-onset primary lateral sclerosis in a chinese family. European Neurology, 2019. 81(1-2): p. 87–93.

17. Yang, Y., et al., Compound heterozygote mutations in SPG7 in a family with adult-onset primary lateral sclerosis. Neurology Genetics, 2016. 2(2).

18. de Boer, E.M., et al., Genetic characterization of primary lateral sclerosis. Journal of Neurology, 2023: p. 1–11.

19. Project MinE: study design and pilot analyses of a large-scale whole-genome sequencing study in amyotrophic lateral sclerosis. European Journal of Human Genetics, 2018. 26(10): p. 1537–1546.

20. Rothstein, J., et al., Answer ALS: A Large-Scale Resource for Sporadic and Familial ALS Combining Clinical Data with Multi-Omics Data from Induced Pluripotent Cell Lines. 2020.

21. Baxi, E.G., et al., Answer ALS, a large-scale resource for sporadic and familial ALS combining clinical and multi-omics data from induced pluripotent cell lines. Nature neuroscience, 2022. 25(2): p. 226–237.

22. Tazelaar, G.H., et al., ATXN1 repeat expansions confer risk for amyotrophic lateral sclerosis and contribute to TDP-43 mislocalization. Brain communications, 2020. 2(2): p. fcaa064.

23. Cooper-Knock, J., et al., Clinico-pathological features in amyotrophic lateral sclerosis with expansions in C9ORF72. Brain, 2012. 135(3): p. 751–764.

24. Blauw, H.M., et al., A large genome scan for rare CNVs in amyotrophic lateral sclerosis. Human molecular genetics, 2010. 19(20): p. 4091–4099.

25. Elden, A.C., et al., Ataxin-2 intermediate-length polyglutamine expansions are associated with increased risk for ALS. Nature, 2010. 466(7310): p. 1069–1075.

26. DeJesus-Hernandez, M., et al., Expanded GGGGCC hexanucleotide repeat in noncoding region of C9ORF72 causes chromosome 9p-linked FTD and ALS. Neuron, 2011. 72(2): p. 245–256.

27. Tazelaar, G.H., et al., Association of NIPA1 repeat expansions with amyotrophic lateral sclerosis in a large international cohort. Neurobiology of aging, 2019. 74: p. 234. e9-234. e15.

28. Mehta, P.R., et al., The impact of age on genetic testing decisions in amyotrophic lateral sclerosis. Brain, 2022. 145(12): p. 4440–4447.

29. Brown, R.H. and A. Al-Chalabi, Amyotrophic lateral sclerosis. New England Journal of Medicine, 2017. 377(2): p. 162–172.

30. Balicza, P., et al., Genetic background of the hereditary spastic paraplegia phenotypes in Hungary—An analysis of 58 probands. Journal of the neurological sciences, 2016. 364: p. 116–121.

31. Arnoldi, A., et al., A clinical, genetic, and biochemical characterization of SPG7 mutations in a large cohort of patients with hereditary spastic paraplegia. Human mutation, 2008. 29(4): p. 522–531.

32. Hehr, U., et al., Long-term course and mutational spectrum of spatacsin-linked spastic paraplegia. Annals of Neurology: Official Journal of the American Neurological Association and the Child Neurology Society, 2007. 62(6): p. 656–665.

33. Stevanin, G., et al., Mutations in SPG11 are frequent in autosomal recessive spastic paraplegia with thin corpus callosum, cognitive decline and lower motor neuron degeneration. Brain, 2008. 131(3): p. 772–784.

34. Roxburgh, R.H., et al., The p. Ala510Val mutation in the SPG7 (paraplegin) gene is the most common mutation causing adult onset neurogenetic disease in patients of British ancestry. Journal of neurology, 2013. 260(5): p. 1286–1294.

35. Yamamoto-Watanabe, Y., et al., A Japanese ALS6 family with mutation R521C in the FUS/TLS gene: a clinical, pathological and genetic report. Journal of the neurological sciences, 2010. 296(1-2): p. 59–63.

36. Kwiatkowski Jr, T., et al., Mutations in the FUS/TLS gene on chromosome 16 cause familial amyotrophic lateral sclerosis. Science, 2009. 323(5918): p. 1205–1208.

37. Chiò, A., et al., A de novo missense mutation of the FUS gene in a “true” sporadic ALS case. Neurobiology of aging, 2011. 32(3): p. 553. e23-553. e26.

38. Nicholson, G., et al., Distinctive genetic and clinical features of CMT4J: a severe neuropathy caused by mutations in the PI (3, 5) P2 phosphatase FIG4. Brain, 2011. 134(7): p. 1959–1971.

39. Raczy, C., et al., Isaac: ultra-fast whole-genome secondary analysis on Illumina sequencing platforms. Bioinformatics, 2013. 29(16): p. 2041–2043.

40. Chang, C.C., et al., Second-generation PLINK: rising to the challenge of larger and richer datasets. Gigascience, 2015. 4(1): p. s13742–015-0047-8.

41. Purcell, S., et al., PLINK: a tool set for whole-genome association and population-based linkage analyses. The American journal of human genetics, 2007. 81(3): p. 559–575.

42. Danecek, P., et al., The variant call format and VCFtools. Bioinformatics, 2011. 27(15): p. 2156–2158.

43. Li, H. and R. Durbin, Fast and accurate short read alignment with Burrows–Wheeler transform. bioinformatics, 2009. 25(14): p. 1754–1760.

44. DePristo, M.A., et al., A framework for variation discovery and genotyping using next-generation DNA sequencing data. Nature genetics, 2011. 43(5): p. 491–498.

45. Van der Auwera, G.A. and B.D. O’Connor, Genomics in the cloud: using Docker, GATK, and WDL in Terra. 2020: O’Reilly Media.

46. Wang, K., M. Li, and H. Hakonarson, ANNOVAR: functional annotation of genetic variants from high-throughput sequencing data. Nucleic acids research, 2010. 38(16): p. e164–e164.

47. McLaren, W., et al., The ensembl variant effect predictor. Genome biology, 2016. 17(1): p. 1–14.

48. Abel, O., et al., ALSoD: A user-friendly online bioinformatics tool for amyotrophic lateral sclerosis genetics. Human mutation, 2012. 33(9): p. 1345–1351.

49. Hinrichs, A.S., et al., The UCSC genome browser database: update 2006. Nucleic acids research, 2006. 34(suppl_1): p. D590–D598.

50. Dolzhenko, E., et al., Detection of long repeat expansions from PCR-free whole-genome sequence data. Genome research, 2017. 27(11): p. 1895–1903.

51. Kaplan, E.L. and P. Meier, Nonparametric estimation from incomplete observations. Journal of the American statistical association, 1958. 53(282): p. 457–481.

52. Therneau, T.M. and T. Lumley, Package ‘survival’. R Top Doc, 2015. 128(10): p. 28–33.

53. Kassambara, A., et al., Package ‘survminer’. Drawing Survival Curves using ‘ggplot2’(R package version 03 1), 2017.

54. Computing, R., R: A language and environment for statistical computing. Vienna: R Core Team, 2013.

