## Supplemental Table 1, 2 for "Genetic and phenotype analyses of primary lateral sclerosis datasets from international cohorts"

**Genetic screening of a large primary lateral sclerosis dataset from international cohorts supplementary tables**

Supplementary Table 1 – Results of genetic testing for Moderate impact variants. The last two columns indicate whether a variant was classified clinically actionable and in a gene present in the corresponding panel. SETX**^*^:** same individual.

| Gene name | Variant | Protein change | chr | POS  (hg19) | REF | ALT | Large gene | Genomics England panel |
| --- | --- | --- | --- | --- | --- | --- | --- | --- |
| *ALS2* | c.1280C>A | p.Thr427Lys | chr2 | 202622316 | G | T | NO | NO |
| *ATXN2* | c.1322C>T | p.Tyr441Met | chr12 | 111948236 | G | A | NO | NO |
| *C9orf72* | c.971A>G | p.His324Arg | chr9 | 27556679 | T | C | NO | NO |
| *ERBB4* | c.1829C>G | p.Pro610Arg | chr2 | 212530090 | G | C | NO | YES |
| *FIG4* | c.1373dup | p.Leu458Ffs*5 | chr6 | 110083391 | A | AT | NO | YES |
| *FIG4* | c.68G>A | p.Arg23Lys | chr6 | 110036282 | G | A | NO | YES |
| *FUS* | c.1561C>T | p.Arg521Cys | chr16 | 31202739 | C | T | YES | YES |
| *MATR3* | c.1640A>G | p.Asn547Ser | chr5 | 138665044 | A | G | YES | YES |
| *SETX** | c.5084A>C | p.Gln1695Pro | chr9 | 135201901 | T | G | NO | YES |
| *SETX** | c.3311A>C | p.Gln1104Pro | chr9 | 135203674 | T | G | NO | YES |
| *SPG7* | c.233T>A | p.Leu78X | chr16 | 89576947 | T | A | NO | NO |
| *SPG7* | c.1529C>T | p.Ala510Val | chr16 | 89613145 | C | T | NO | NO |
| *SPG11* | c.1951C>T | p.Arg651X | chr15 | 44920983 | G | A | NO | YES |
| *SQSTM1* | c.C1175C>T | p.Pro392Leu | chr5 | 179263445 | C | T | YES | YES |
| *TARDBP* | c.71G>A | p.Gly24Asp | chr1 | 11073855 | G | A | YES | YES |
| *TBK1* | c.2044delA | p.T682Hfs*2 | chr12 | 64891511 | CA | C | YES | YES |

Supplementary Table 2 - Results of genetic testing for High impact variants. The last two columns indicate whether a variant was classified clinically actionable and in a gene present in the corresponding panel. PFN1**^*^:** same individual.

| Gene name | Variant | Protein change | Chr | POS  (hg19) | REF | ALT | Large gene | Genomics England panel |
| --- | --- | --- | --- | --- | --- | --- | --- | --- |
| *ALS2* | c.C1115G | p.Pro372Arg | chr2 | 202622481 | G | C | NO | NO |
| *ANAX11* | c.T1010A | p.Leu337His | chr10 | 81923309 | A | T | NO | YES |
| *PFN1** | c.A351T | p.Glu117Asp | chr17 | 4849267 | T | A | YES | YES |
| *PFN1** | c.A350G | p.Glu117Gly | chr17 | 4849268 | T | C | YES | YES |
| *PNPLA6* | c.350A>G | p.Glu117Gly | chr19 | 7625651 | G | A | NO | NO |
| *LIPC* | c.1214C>T | p.Thr405Met | chr15 | 58855748 | C | T | NO | NO |
| *SPAST* | c.134C>A | p.Pro45Gln | chr2 | 32289034 | C | A | NO | NO |
| *SPG11* | c.5121G>T | p.Glu1707Asp | chr15 | 44877834 | C | A | NO | NO |
| *SQSTM1* | c.961C>T | p.Arg321Cys | chr5 | 179260238 | C | T | NO | NO |
| *VAPB* | c.474_476del | p.S160del | chr20 | 57016039 | GTTC | G | YES | YES |
